# Age- and Sex-Specific Norms for ADHD Symptoms and Performance on the Vanderbilt Assessment Scale

**DOI:** 10.64898/2026.10.12.26365313

**Authors:** Steven W. Evans, Shreya Kupur Bhaskar, Yinuo Liu, Eric A. Youngstrom, Julie Sarno Owens, Emily A. Rosenthal

## Abstract

**Objective**: To provide national norms, percentiles, and results of factor analyses for the ADHD symptoms and performance items on the parent-report and self-report versions of the Vanderbilt Assessment Scale (VAS) for youth ages 6 to 17. We hypothesized that symptom items would demonstrate separate inattention and hyperactivity/impulsivity factors, performance items would demonstrate interpretable latent dimensions, measurement invariance would be supported across age and sex, and symptom ratings would vary systematically by age and sex.

**Method**: In a nationally stratified survey conducted in 2025, 2,230 parents of youth aged 6-17 years and 996 adolescents aged 12-17 years completed Vanderbilt Assessment Scale items. Both versions asked about the frequency of 18 ADHD symptoms and performance in 8 domains. To evaluate the psychometric properties and age-related normative curves of the VAS across parent- and youth-report forms, we conducted exploratory factor analysis, confirmatory factor analysis, and measurement invariance testing. We also generated age and sex-specific norms.

**Results:** Factor analyses produced the expected two symptom factors and two performance factors. Developmental and sex-specific patterns of ratings of inattention and hyperactivity/impulsivity symptoms indicated that after increasing slightly during the preadolescent years, parent ratings of inattention symptoms declined and remained stable. In contrast, ratings of hyperactivity/impulsivity declined steadily with age. Normative data were generated using GAMLSS estimated centiles from fitted probability distributions.

**Conclusions**: This study provides norms for the parent-report and adolescent self-report VAS. Findings underscore the importance of using sex- and age-based norms to better inform estimates of severity and diagnostic decision making.

## Introduction

Attention Deficit Hyperactivity Disorder (ADHD) is a common neurodevelopmental disorder associated with substantial academic, social, and family impairment and elevated risk for adverse long-term outcomes.^1–2^ Accurate assessment across development is critical for guiding treatment as well as prevention of associated debilitating outcomes.^3^

There are a variety of psychometrically sound rating scales used to assess ADHD symptoms and impairment in youth, but many lack national norms and other psychometric evaluations.^4^ Normative data on ratings of symptoms have been reported for the Disruptive Behavior Disorders Rating Scale^5^; however, these are limited to teacher ratings of children aged 14 and younger. Other scales include norms by age and sex for parent and teacher ratings into adolescence^6^; however, normative data for self-ratings are not available. Most measures of ADHD symptoms consist of ratings of the 18 symptoms of the disorder from the Diagnostic and Statistical Manual of Mental Disorders (DSM), with a symptom considered present when the rater endorses it occurs ’often’ or ’very often’ (or ‘pretty much’ or ‘very much’) over the last six months. For youth younger than 17 years of age, the endorsement of six or more symptoms of inattention (9 items) or six or more symptoms of hyperactivity/impulsivity (9 items) meets the symptom criteria for at least one of the three possible presentations of ADHD.^7^ To establish an ADHD diagnosis, other criteria must also be met, including the onset of several symptoms prior to age 12, the presence of the symptoms in two or more settings, and symptom-related impairment.

The Vanderbilt Assessment Scale (VAS) is widely used in clinical settings, and prior studies support the reliability and validity of its ADHD symptom items.^8–10^ Anderson and colleagues provided U.S. national norms and percentiles for parent ratings of ADHD symptoms for children ages 5 – 12 with a nationally stratified survey sample of 1,570 caregivers.^11^ However, norms are not available for parent- and self-reported symptoms for adolescents. As it is often difficult to obtain ratings from secondary school teachers, this limitation is clinically meaningful, as parent and self-report are often the only feasible options for gathering symptom ratings in routine clinical practice. Further, there has been limited evaluation of the eight performance items, though some evidence supports the concurrent validity of these scores with other functioning scales^12^ and their utility in identifying comorbid learning disabilities.^13^ However, prior studies have not explored the factor structure of the performance items nor reported national norms. Therefore, it is unclear whether these items are appropriate to be used to determine the DSM-V impairment criterion for diagnosing ADHD.

We hypothesized that symptom items would demonstrate separate inattention and hyperactivity/impulsivity factors, performance items would demonstrate interpretable academic and social dimensions, measurement invariance would be supported across age and sex, and symptom ratings would vary systematically as a function of age and sex. We also examined the percentage of the sample meeting symptom-criteria for ADHD (≥6 hyperactive/impulsive or ≥6 inattentive symptoms) by age bands and sex. These results enhance the utility of the VAS for assessing ADHD in both clinical and research contexts.

## Method

### Eligibility Criteria

Eligible participants were parents or legal guardians of a child aged 6-17 years residing in the United States. Adolescent self-report participants were youth aged 12-17 years whose parents also participated.

### Participants

This study was approved by the Institutional Review Board at [masked]. The survey agency YouGov was used to recruit and administer questionnaires to parents and adolescents in the United States. These questionnaires included the VAS ADHD symptom and performance items. YouGov is a research and data analytics company that has a global panel of over 29 million registered members with a significant portion in the U.S., which enables the company to conduct national surveys. Parents of children and teens aged 6-17 (*n*=2,230) and adolescents aged 12-17 (*n*=996) completed the VAS. YouGov collected and cleaned all responses and applied stratification procedures based on key demographics (child sex, race, ethnicity, and parent education level). Survey responses were screened for quality prior to inclusion resulting in a nationally stratified U.S. sample, and demographic characteristics of the sample are reported in Table 1.

**Table 1.** Participant demographic characteristics

|  | **Parent Sample**<br>**(n=2,230)** | **Adolescent Sample**<br>**(n=996)** |
| --- | --- | --- |
| **Child age** | 11.7 (3.40) | 14.4 (1.6) |
| 6-7 years | 15.6% | - |
| 8-9 years | 14.8% | - |
| 10-11 years | 14.4% | - |
| 12-13 years | 19.5% | 34.3% |
| 14-15 years | 20% | 35.5% |
| 16-17 years | 15.6% | 30.1% |
| **Child sex** |  |  |
| Female | 47.4% | 49.5% |
| Male | 52.6% | 50.5% |
| **Race** |  |  |
| White | 75.4% | 74.8% |
| Black or African American | 8.8% | 9.7% |
| Asian | 3.7% | 1.8% |
| American Indian or Alaska Native | 1.7% | 0.7% |
| Native Hawaiian or Pacific Islander | 0.3% | 0.2% |
| Some other race | 4.7% | 3.3% |
| Multiple races selected | 4.0% | 8.9% |
| **Ethnicity** |  |  |
| Hispanic, Latino, or Spanish Origin | 15.7% | 10.9% |
| Not Hispanic, Latino, or Spanish Origin | 80.9% | 85.6% |
| **Parent/guardian education** |  |  |
| High school graduate or higher | 50% | 48.7% |
| Bachelor's degree or higher | 47.6% | 48.8% |
| **Median Household Income** | \$70,000-\$79,999 | \$70,000-\$79,999 |

### Study Design

This was a cross-sectional observational psychometric and normative study using a nationally stratified survey sample of U.S. parents and adolescents. Data collection occurred between July 2025 and March 2026 through the YouGov online survey platform.

### Measures

Parents and adolescents completed the 18 VAS ADHD symptom items and the eight performance items assessing school, academic, relational, and activity functioning. Symptom items were rated as occurring 0 (*never*), 1 (*occasionally*), 2 (*often*), or 3 (*very often*). Consistent with Vanderbilt scoring procedures, symptom items rated as 2 (“often”) or 3 (“very often”) were considered positive symptom endorsements for symptom-count analyses. Performance items were rated as 1 (*excellent*), 2 (*above average*), 3 (*average*), 4 (*somewhat of a problem*), and 5 (*problematic*). Performance items rated as 4 (“Somewhat of a problem”) or 5 (“Problematic”) were considered positive performance endorsements (i.e., indicative of significant concerns) for performance-count analyses. Parent and self-report versions of the VAS are freely available from the National Institute for Children’s Health Quality website.^14^

### Analyses

All analyses were conducted in R (v4.5.2) using RStudio (v2026.01.1). Exploratory factor analysis (EFA), confirmatory factor analysis (CFA), measurement invariance testing, and regression-based continuous norming were conducted to evaluate the psychometric and cross-sectional age properties of the VAS across parent- and adolescent-report forms. Ordinal item responses were analyzed using polychoric correlations. Symptom responses were collapsed into three ordinal categories (0,1, and a combined category of 2 and 3), and invalid or skipped responses were treated as missing.

The parent sample was randomly divided into exploratory (n = 1,115) and confirmatory (n = 1,115) subsamples stratified by age group. The adolescent sample was divided into exploratory (n = 499) and confirmatory (n = 497). Both parent and adolescent subsamples were stratified by age group (6-11 and 12-17 years for the parent-report sample; 12-14 and 15-17 years for the adolescent-report sample) to ensure cross-validation of factor structures. EFA was conducted using minimum residual estimation with oblimin rotation, which allowed for correlated factors because ADHD symptom domains were expected to be related. Factor retention was based on parallel analysis, supported by scree plots and eigenvalues. CFA models were specified with Inattention and Hyperactivity/Impulsivity factors for symptom items and Academic and Social factors for performance items.

CFA and measurement invariance analyses were conducted with the confirmatory subsamples using weighted least squares means and variance (WLSMV) for ordered categorical indicators. Model fit was evaluated using the comparative fit index (CFI), root mean square error of approximation (RMSEA), and standardized root mean square residual (SRMR). Measurement invariance across age group and sex was evaluated using sequential multi-group CFA models testing configural, metric, and scalar invariance. Changes in CFI (ΔCFI) were used as the primary criterion for evaluating invariance across nested models, with ΔCFI ≤ 0.01 indicating support for invariance. Factor analyses were conducted using recoded symptom responses (0, 1, and combined 2/3 categories) to improve stability of ordinal analyses. Continuous norming analyses retained the original dimensional scale scores and modeled age- and sex-specific distributions of summed symptom and performance scores.

Based on the results of the factor analyses, four domain scores were used to calculate centiles. The Inattention score was calculated as the sum of the nine DSM-based inattention symptom items (possible range = 0-27). The Hyperactivity/Impulsivity score was calculated as the sum of the nine hyperactivity/impulsivity symptom items (possible range = 0-27). For performance items, Academic Performance scores were calculated by summing ratings of overall school performance, reading, writing, and mathematics (possible range = 4-20). Social Performance scores were calculated by summing ratings of relationships with parents, siblings, peers, and participation in organized activities (possible range = 4-20). Higher scores indicated greater symptom severity or functional difficulty.

Age-related normative curves were estimated using generalized additive models for location, scale, and shape (GAMLSS).^15–16^ This method was chosen because GAMLSS allow normative percentile curves to change continuously with age rather than relying on broad age categories. Because Vanderbilt symptom scores are discrete, positively skewed, and included a high proportion of zero symptom endorsements, candidate non-normal distributions were evaluated within the GAMLSS framework. Within each model, the outcome distribution was modeled as a function of a sex indicator, a penalized spline for age, and a sex-specific penalized spline for age, allowing age-related trajectories to differ by sex. Candidate distributions were compared using the Bayesian information criteria (BIC), worm plots, and Q-statistics. Final models used zero-inflated negative binomial (ZINBI) for inattention symptom domain, negative binomial (NBI) distributions for hyperactivity symptom domain, and beta-binomial (BB) distributions for academic and social performance domains. Age- and sex-specific percentile curves (10^th^-95^th^ percentiles) were generated from the final fitted models for parent-reported and adolescent-reported symptom and performance domains. Missing item responses were handled using pairwise-present correlations in EFA and WLSMV estimation procedures in CFA.

The prevalence of individuals meeting ADHD symptom thresholds was estimated using DSM-aligned thresholds (≥6 symptoms for inattention and/or hyperactivity/impulsivity) with 95% confidence intervals.

## Results

### Exploratory Factor Analysis

EFA supported a stable two-factor structure for parent ratings of both symptom and performance domains. Symptom items clustered into the expected Inattention and Hyperactivity/Impulsivity factors with minimal cross-loadings, supporting clear differentiation between symptom dimensions (see Table 2). Performance items also demonstrated a coherent two-factor structure (see Table S2) clustering into an Academic factor and a Social factor.

**Table 2.** Exploratory factor analysis of parent and adolescent self-ratings of symptoms

| Symptom | Parent Ratings (n=1,115) |  | Adolescent-Ratings (n=499) |  |
| --- | --- | --- | --- | --- |
|  | Inattention | Hyperactivity | Inattention | Hyperactivity |
| 1. Does not pay attention to details or makes careless mistakes with, for example, homework | **0.86** | -0.06 | **0.94** | -0.17 |
| 2. Has difficulty keeping attention to what needs to be done | **0.87** | 0.05 | **0.87** | 0.04 |
| 3. Does not seem to listen when spoken to directly | **0.62** | 0.26 | **0.71** | 0.14 |
| 4. Does not follow through when given directions and fails to finish activities (not due to refusal or failure to understand) | **0.89** | -0.02 | **0.85** | 0.01 |
| 5. Has difficulty organizing tasks and activities | **0.97** | -0.11 | **0.92** | -0.05 |
| 6. Avoids, dislikes, or does not want to start tasks that require ongoing mental effort | **0.85** | 0.00 | **0.78** | 0.02 |
| 7. Loses things necessary for tasks or activities (toys, assignments, pencils, or books) | **0.71** | 0.13 | **0.68** | 0.13 |
| 8. Is easily distracted by noises or other stimuli | **0.61** | 0.26 | **0.71** | 0.10 |
| 9. Is forgetful in daily activities | **0.84** | 0.04 | **0.76** | 0.11 |
| 10. Fidgets with hands or feet or squirms in seat | 0.33 | **0.53** | 0.44 | 0.38 |
| 11. Leaves seat when remaining seated is expected | 0.07 | **0.82** | 0.04 | **0.79** |
| 12. Runs about or climbs too much when remaining seated is expected | -0.06 | **0.93** | -0.09 | **0.95** |
| 13. Has difficulty playing or beginning quiet play activities | 0.05 | **0.79** | 0.22 | **0.64** |
| 14. Is “on the go” or often acts as if “driven by a motor” | -0.09 | **0.89** | 0.09 | **0.74** |
| 15. Talks too much | -0.03 | **0.73** | 0.06 | **0.63** |
| 16. Blurts out answers before questions have been completed | 0.06 | **0.77** | 0.00 | **0.85** |
| 17. Has difficulty waiting his or her turn | 0.03 | **0.83** | -0.01 | **0.87** |
| 18. Interrupts or intrudes in on others’ conversations and/or activities | 0.10 | **0.76** | 0.04 | **0.77** |

Notes: Symptoms 1 – 9: Inattention factor; Symptoms 10-18: Hyperactivity/Impulsivity factor. Factor extraction was performed using minimum residual (minres) method with oblimin rotation. Larger factor loadings indicate stronger association between items and latent factors.

Analysis of adolescent self-reports replicated the same two-factor structure observed in parent reported data. Symptom items loaded strongly onto Inattention and Hyperactivity/Impulsivity factors except for item 10 (fidgets; see Table 2), and performance items loaded onto coherent Academic and Social factors (see Table S2).

In practical terms, symptom items grouped into the expected Inattention and Hyperactivity/Impulsivity domains and performance items grouped into Academic and Social functioning domains, supporting the conceptual organization of the Vanderbilt Assessment Scale.

### Confirmatory Factor Analysis

CFA also supported the proposed two-factor structure parent- and adolescent-report symptom and performance models. Parent-report symptom models demonstrated good fit (CFI = 0.97, RMSEA = 0.08, SRMR = 0.05), and parent-report performance models demonstrated strong fit (CFI = 0.99, RMSEA = 0.06, SRMR = 0.05).

Models of adolescent-reported symptoms also demonstrated good fit (CFI = 0.98, RMSEA = 0.08, SRMR = 0.06) and supported the proposed two-factor structure of inattention and hyperactivity/Impulsivity. In contrast, adolescent-report performance models demonstrated weaker absolute fit (CFI = 0.97, RMSEA = 0.10, SRMR = 0.10). Although the CFI indicated acceptable comparative fit, the elevated RMSEA and SRMR suggest that the two-factor performance model fit less well.

In summary, these findings indicate that items on the VAS consistently measured the intended symptom and performance domains for both parent- and adolescent-report forms.

### Measurement Invariance

Measurement invariance testing supported configural, metric, and scalar invariance across both age group and sex for parent- and adolescent-report symptom and performance models. Parent-reported symptom models demonstrated good configural fit across age (CFI= 0.98, RMSEA= 0.07, SRMR= 0.05) and sex (CFI= 0.97, RMSEA= 0.08, SRMR= 0.06) (see Table S1). Changes in fit across metric and scalar models were minimal (all ΔCFI ≤ 0.008) and below the recommended thresholds that would indicate non-invariance (ΔCFI < 0.01 and ΔRMSEA < 0.015^17–18^), supporting full measurement invariance. Parent-report performance models showed a similar pattern, with good configural fit and negligible changes across nested models (ΔCFI ≤ 0.002), providing evidence for full measurement invariance (see Table S3). In practical terms, these findings suggest that Vanderbilt scores may be interpreted similarly across age and sex groups.

Adolescent-report symptom models likewise demonstrated good configural fit across age (CFI = 0.98, RMSEA = 0.07, SRMR = 0.07) and sex (CFI = 0.98, RMSEA = 0.07, SRMR = 0.07), with minimal changes across metric and scalar models (all ΔCFI ≤0.005) (see Table S1). Although the adolescent-report performance model showed weaker absolute fit, changes in fit across nested models remained minimal (ΔCFI ≤ 0.006), supporting measurement invariance across age and sex (see Table S3).

Overall, results from both parent- and adolescent-report models support the validity of comparisons across age and sex. Therefore, differences in Vanderbilt scores observed across age and sex groups are likely to reflect true differences in symptom levels rather than differences in how items function across groups.

### Age and Sex Specific Trajectories

Age- and sex-specific percentile trajectories from the GAMLSS models demonstrated distinct patterns across symptom and performance domains (see Figures 1&2; Supplementary Figures S1-S2). Clinically, this means that the same raw Vanderbilt score may represent average symptom levels at one age but elevated symptom levels at another age. Age-specific norms therefore provide a more meaningful interpretation than a single cut score applied across development.

**Figure 1.**
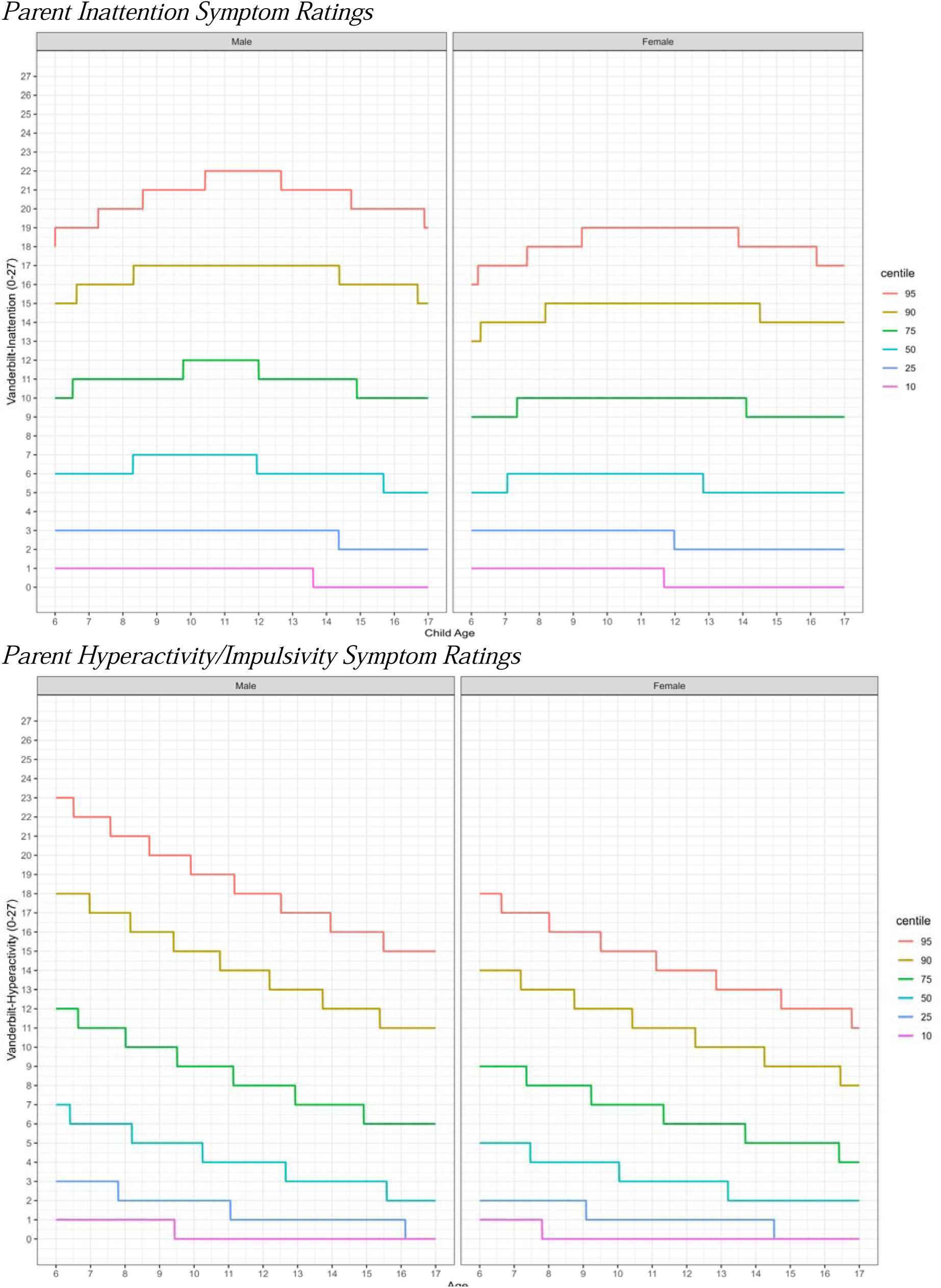
Parent Inattention Symptom Ratings Parent Hyperactivity/Impulsivity Symptom Ratings

**Figure 2.**
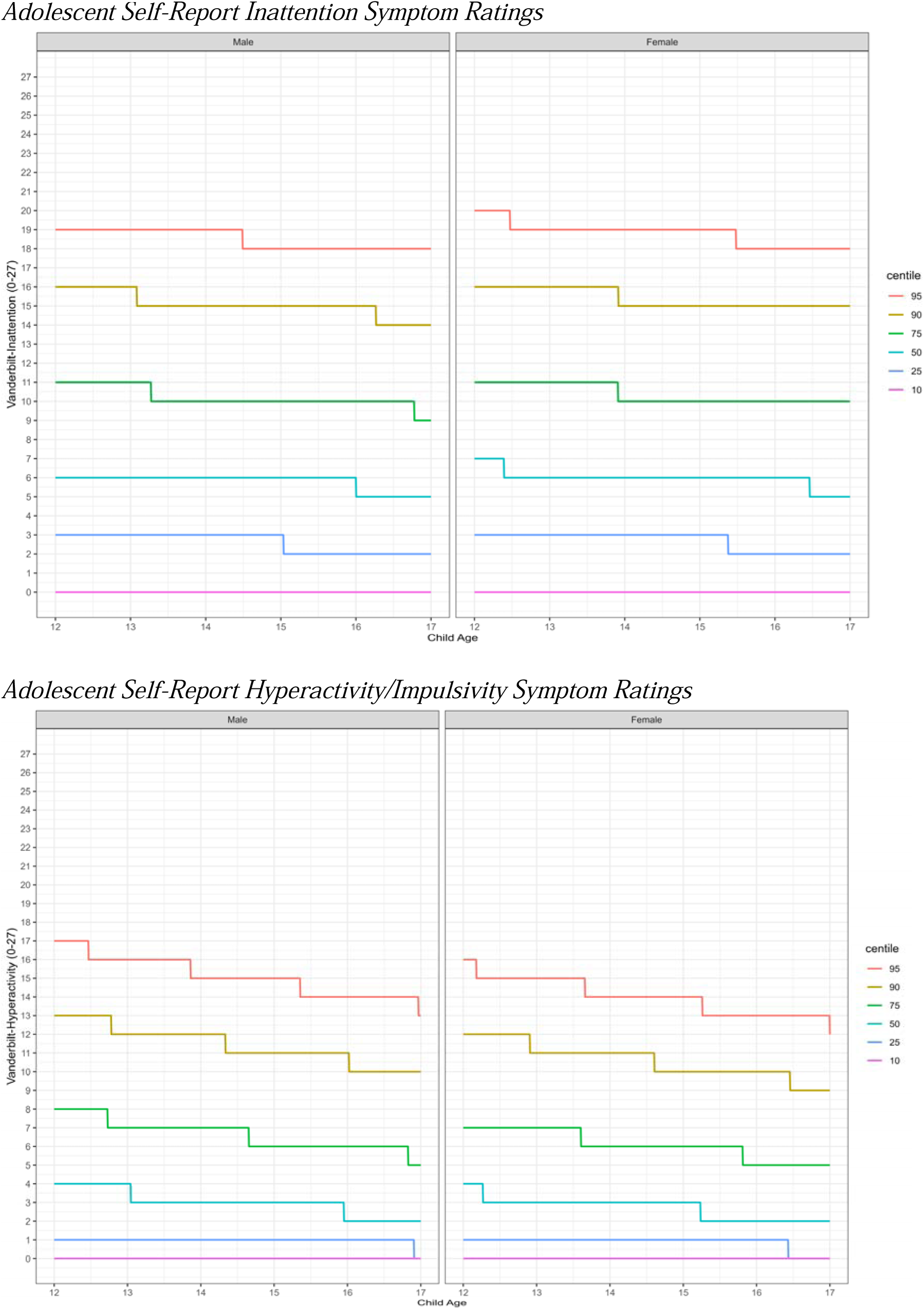
Adolescent Self-Report Inattention Symptom Ratings Adolescent Self-Report Hyperactivity/Impulsivity Symptom Ratings

Inattention trajectories varied by informant. Across sex, parent-reported inattention showed a nonlinear pattern, with increasing scores during childhood, peak levels in early adolescence, and subsequent decline through late adolescence. Self-reported inattention remained relatively stable across adolescence, with minimal age-related differences and similar trajectories between sexes.

Hyperactivity/Impulsivity showed an age-related decline across all percentiles in both parent- and adolescent-report measures. The decline was most pronounced at higher symptom percentiles, with males demonstrating slightly higher trajectories than females across comparable ages.

Performance domains showed substantially greater stability than symptoms across development. Academic performance trajectories remained largely consistent across childhood and adolescence for both parent and adolescent reports, with only minor variation across percentiles. Social performance showed modest differences across ages with overall stable trajectories across age and sex. Given that ratings were anchored by raters’ perceptions of ‘average’ (3 = *average*) flat profiles were expected as the definition of average changes with age.

Although factor analyses supported distinct Academic and Social domains, the adolescent self-report performance model demonstrated weaker fit than the symptom models. Therefore, adolescent self-reported performance ratings should be interpreted somewhat more cautiously than symptom ratings, particularly when making comparisons across individuals.

### ADHD Symptom Prevalence

Prevalence estimates of youth with ADHD symptoms meeting DSM-aligned symptom thresholds (≥6 symptoms) per parent-report showed expected developmental and sex-related patterns (see Figure 3). The prevalence of youth meeting inattention symptom thresholds ranged from 10.2%-19% in males and 4.7%-13.8% in females, with highest prevalence observed during early to mid-adolescence. The prevalence of youth meeting hyperactivity/impulsivity symptom thresholds declined with age in both sexes, ranging from 2.4%-15.6% in males and 2.2%-8.3% in females with the highest prevalence observed in youth ages 8-9.

**Figure 3.**
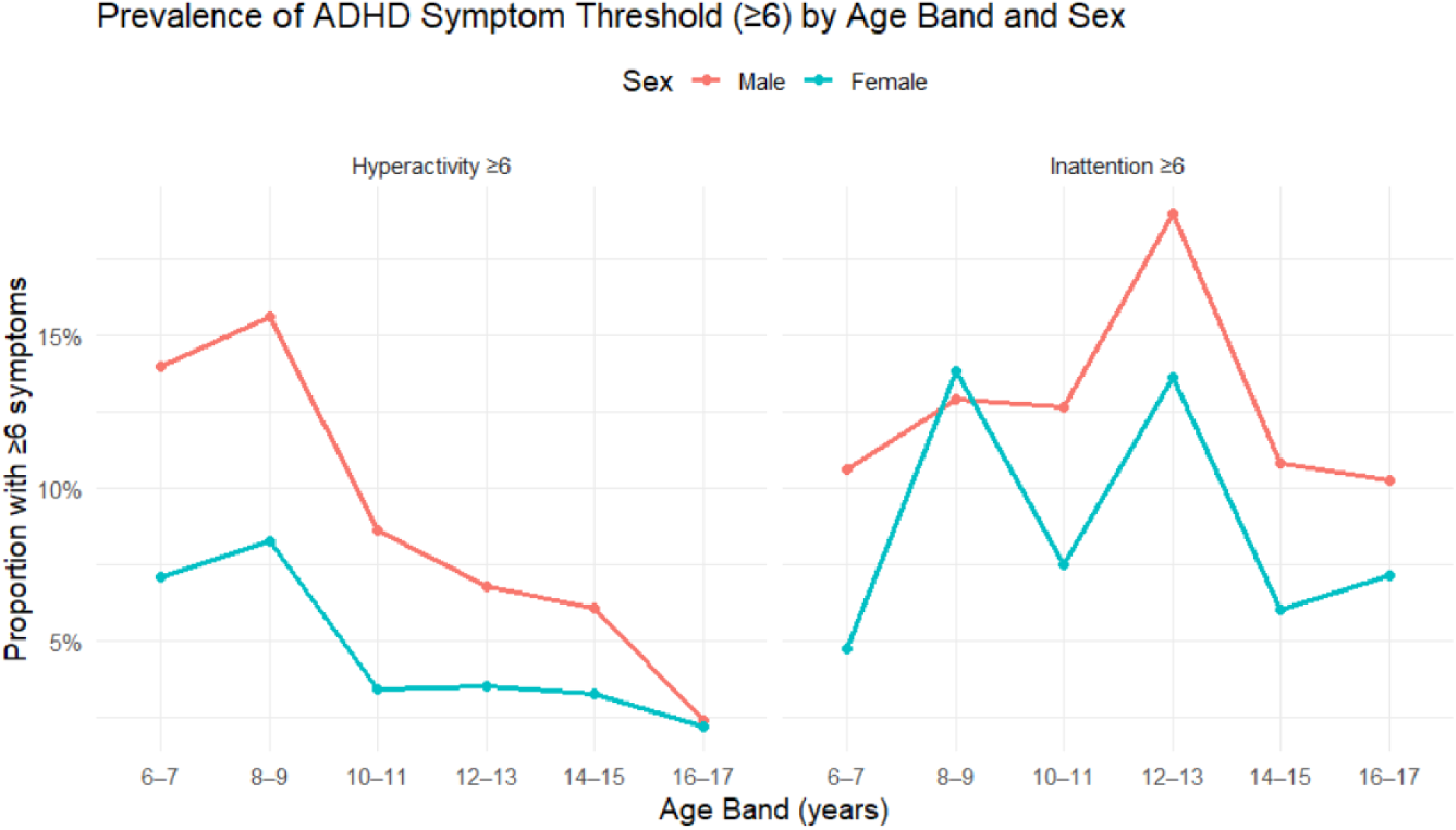
Proportion of Sample Based on Parent Ratings

## Discussion

### Symptoms

The results of the EFA and CFA revealed a two-factor structure of the VAS ADHD symptom items, with an inattention dimension and a hyperactive/impulsive dimension that are invariant to sex and age. This replicates other studies conducted with large samples of parents of elementary school-aged children and adolescents.^6,19^ The “fidgets” item had the lowest Hyperactivity/Impulsivity loading for both parent and adolescent ratings. Because the reason for this difference cannot be determined from the present data, replication and item-level investigation are needed before drawing conclusions about how this behavior is interpreted.

Our findings also highlight the importance of considering developmental level and sex when assessing the severity of ADHD symptoms. Consistent with other studies,^6^ parent ratings of hyperactivity/impulsivity decline with child age. Uniquely, our results reveal that a similar pattern exists for adolescent self-report. Thus, when interpreting the severity of symptoms, it is critical to compare a child’s scores to the normative data for that age. For example, at age 6, a parent-rated score of 12 on the hyperactive/impulsive subscale is fairly normative (within the 75^th^ percentile); yet this same score at age 16 or 17 would place the child in the 90^th^ percentile. Consistent with previous studies, both parent and adolescent ratings indicated greater problems for males than females at all ages.^6,19^ Therefore, the developmental atypicality/clinical interpretation of the same level of hyperactive/impulsive symptoms may differ depending on the sex of the child.

In contrast, symptoms of inattention did not decline with age. In fact, parent ratings of inattention increased slightly from childhood to early adolescence and then declined in late adolescence. This pattern of elevated inattention near the onset of puberty and entry to middle school has been reported elsewhere^6,20^ and may be due to physiological changes associated with puberty and the expectation for adolescents to function with increasing independence, particularly in the school setting. To the extent that increases in demands for independent functioning outpace a child’s ability to meet these demands, problems will appear increasingly severe.

The present norms were broadly consistent with earlier studies, although some differences appeared at the 90th percentile.^6,11,21^ These differences may reflect sampling or analytic methods. Unlike other studies that reported norms based on discrete age bands, our use of GAMLSS allowed the distribution of Vanderbilt scores to vary smoothly as a function of age. Consequently, the GAMLSS-derived centiles provide age-specific estimates that change gradually across development rather than exhibiting stepwise changes at arbitrary age-band boundaries. Furthermore, GAMLSS estimates centiles from fitted probability distributions while simultaneously modeling age-related changes in the location and dispersion of the score distribution. By borrowing information across adjacent ages, this approach reduces sampling variability and produces stable normative reference curves rather than empirical percentile estimation within discrete age bands. This offers greater confidence for practitioners when making age-based interpretations and may account for differing norms relative to past studies.

The proportion of youth who exceeded the six-symptom threshold was higher than published estimates of ADHD prevalence.^22^ This was expected, as the present analyses did not assess symptom onset, duration, presentation across settings, impairment, or alternative explanations. However, age and sex differences in the percentage of youth meeting symptom thresholds suggest that determining a diagnosis may be best informed by considering both symptom cutoffs for ADHD and normative levels of symptoms. Future research is needed before age- or sex-specific normative diagnostic thresholds can be recommended.

## Performance

The VAS includes items assessing rater perceptions of performance. This is the first study to evaluate the factor structure and provide normative data on performance ratings completed by parents (ages 6 –17) and adolescents (ages 12-17). Performance items demonstrated an interpretable Academic and Social factor structure particularly for parent report. However, these items assess broad domains of functioning that are influenced by numerous developmental, educational, psychiatric, and environmental factors beyond ADHD. For example, there are many reasons why a child may be exhibiting problems with “Reading”, “Writing”, and “Mathematics” that are unrelated to ADHD; in fact, these items have been found useful for screening for learning disabilities^13^. Consequently, these items should be interpreted as broad indicators of overall functioning rather than ADHD-specific impairment and should be supplemented with clinical interviews and comprehensive measures of functional impairment when making diagnostic decisions about ADHD. In addition, the VAS only offers raters two indicators of impairment (*somewhat of a problem* and *problematic*). In contrast, other measures of impairment offer opportunities to rate a wide range of functioning, with scaling anchors ranging from 4 levels (*no problem* to *very big problem)* on the Columbia Impairment Rating Scale^23^ to 7 levels (*no problem/does not need treatment* to *extreme problem/does need treatment)* on the Impairment Rating Scale^24–25^. Having only two ratings indicating impairment reduces measurement specificity and sensitivity to change when trying to assess response to treatment.

## Implications for use

The normative data provided in this manuscript offer practitioners updated and unique information for assessing the severity of ADHD symptoms as reported by parents for children ages 6 to 17 and as reported by adolescents ages 12-17. The centile figures (see Figures 1 & 2) are provided in a large easy-to-read form as Figures S3 - S6 for use by practitioners.

It is important to note that the difference between diagnostic symptom thresholds and severity interpretations as well as the various ways a child can meet the diagnostic symptom threshold. Namely, a child may meet the 6-symptom diagnostic threshold because 6 symptoms of inattention are rated as occurring “often” (rating of 2) or as “very often” (rating of 3). These differences in ratings would result in different dimensional scores (e.g., 12 vs. 18); and thus, different estimates of severity. For example, for a 6-year-old boy, these two scores equate to different percentile ranks, but neither of them are above the 93^rd^ percentile in severity despite meeting the DSM symptom count threshold. There are multiple possible combinations of ratings that could lead to a child meeting the DSM symptom count criterion, the 93^rd^ percentile, both or neither. Thus, although they can each inform diagnostic decision-making, neither the percentiles nor the symptom counts may be optimal to make a diagnosis on their own. Clinicians should consider other characteristics such as sleep, trauma, comorbidities, and relative onset of various symptom presentations when making diagnostic decisions. Attention problems are one of the most common symptoms in the DSM so the presence of these symptoms does little to help differentiate ADHD from other problems with distinct underlying causes. Further, and beyond the scope of this article, is the importance of also considering the impairment criteria. Indeed, most studies show much lower prevalence rates of ADHD when the impairment criterion is considered in diagnostic decision making.

## Limitations

Our findings should be interpreted in light of possible selection and coverage bias in the online panel, the cross-sectional design, reliance on a single assessment occasion, absence of teacher ratings and diagnostic validation, and limited prior validation of the adolescent self-report form of the VAS. Some of these concerns are mitigated by the large sample size. Future longitudinal and criterion-validation studies should (a) examine how symptom scores relate to independently assessed ADHD diagnosis and (b) identify a valid and useful measure of impairment that, when administered alongside the VAS, can help clinicians assess both ADHD symptoms and impairment.

## Supporting information

Supplementary Tables, Figures and Handouts

## Data Availability

All data produced in the present study are available upon reasonable request to the authors.

## Disclosure

Dr. Evans is the Editor-in-Chief of the journal *School Mental Health* and receives royalties from Springer, Oxford Press, and Taylor & Francis publishing. Dr. Youngstrom is the co-founder and Executive Director of Helping Give Away Psychological Science, a 501c3; he received royalties from the American Psychological Association and Guilford Press, and he holds equity in Joe Startup Technologies.

The data that support the findings of this study are available from the corresponding author, SWE, upon reasonable request subject to institutional policies and data-use agreements.

