## Supplementary Tables, Figures and Handouts for "Age- and Sex-Specific Norms for ADHD Symptoms and Performance on the Vanderbilt Assessment Scale"

**Table S1.** *Confirmatory factor analysis for parent and adolescent self-ratings of symptoms and measurement invariance across age and sex*

| **Parent Ratings (n=1,115)** | | | | | | |
| --- | --- | --- | --- | --- | --- | --- |
| **Group** | **Model** | **CFI** | **RMSEA** | **SRMR** | **ΔCFI** | **ΔRMSEA** |
| Age | Configural | 0.98 | 0.07 | 0.05 | - | - |
| Age | Metric | 0.98 | 0.06 | 0.06 | 0.01 | -0.01 |
| Age | Scalar | 0.98 | 0.07 | 0.05 | -0.01 | 0.01 |
| Sex | Configural | 0.97 | 0.08 | 0.06 | - | - |
| Sex | Metric | 0.98 | 0.06 | 0.06 | 0.01 | -0.01 |
| Sex | Scalar | 0.97 | 0.08 | 0.06 | -0.01 | 0.01 |
| **Adolescent-Ratings (n=497)** | | | | | | |
| **Group** | **Model** | **CFI** | **RMSEA** | **SRMR** | **ΔCFI** | **ΔRMSEA** |
| Age | Configural | 0.98 | 0.07 | 0.07 | - | - |
| Age | Metric | 0.98 | 0.06 | 0.07 | 0.01 | -0.01 |
| Age | Scalar | 0.98 | 0.07 | 0.07 | -0.01 | 0.01 |
| Sex | Configural | 0.98 | 0.07 | 0.07 | - | - |
| Sex | Metric | 0.98 | 0.06 | 0.07 | 0.01 | -0.01 |
| Sex | Scalar | 0.98 | 0.07 | 0.07 | -0.01 | 0.01 |

Note: CFI values ≥ .95, RMSEA ≤ .08, and SRMR ≤ .08 generally indicate acceptable model fit.

**Table S2.** *Exploratory factor analysis of parent and adolescent self-ratings of performance*

|  | **Parent Ratings (n=1,115)** | | **Adolescent-Ratings**  **(n=499)** | |
| --- | --- | --- | --- | --- |
| **Performance Items** | **Academic** | **Social** | **Academic** | **Social** |
| Overall School Performance | **0.89** | -0.06 | **0.81** | 0.11 |
| Reading | **0.93** | -0.01 | **0.93** | -0.05 |
| Writing | **0.81** | 0.13 | **0.87** | -0.03 |
| Mathematics | **0.64** | -0.01 | **0.72** | 0.02 |
| Relationship with Parents | 0.15 | **0.84** | 0.15 | **0.72** |
| Relationship with Siblings | -0.09 | **0.92** | -0.07 | **0.89** |
| Relationship with Peers | -0.03 | **0.90** | -0.04 | **0.93** |
| Participation in organized activities | 0.08 | **0.66** | 0.16 | **0.63** |

Notes: Factor extraction was performed using minimum residual (minres) method with oblimin rotation. Larger factor loadings indicate stronger association between items and latent factors.

**Table S3.** *Confirmatory factor analysis for parent and adolescent ratings of performance and measurement invariance across age and sex*

| **Parent-Ratings (n=1,115)** | | | | | | | | | | | |
| --- | --- | --- | --- | --- | --- | --- | --- | --- | --- | --- | --- |
| **Group** | **Model** | | **CFI** | | **RMSEA** | | **SRMR** | | **ΔCFI** | | **ΔRMSEA** |
| Age | Configural | | 0.99 | | 0.05 | | 0.05 | | - | | - |
| Age | Metric | | 0.99 | | 0.05 | | 0.05 | | 0.00 | | 0.00 |
| Age | Scalar | | 0.99 | | 0.05 | | 0.05 | | 0.00 | | 0.00 |
| Sex | Configural | | 0.99 | | 0.06 | | 0.06 | | - | | - |
| Sex | Metric | | 0.99 | | 0.05 | | 0.06 | | 0.00 | | 0.00 |
| Sex | Scalar | | 0.99 | | 0.05 | | 0.06 | | 0.00 | | 0.00 |
| **Adolescent-Ratings (n=497)** | | | | | | | | | | | |
| **Group** | | **Model** | | **CFI** | | **RMSEA** | | **SRMR** | | **ΔCFI** | **ΔRMSEA** |
| Age | | Configural | | 0.97 | | 0.10 | | 0.12 | | - | - |
| Age | | Metric | | 0.97 | | 0.08 | | 0.12 | | 0.01 | -0.02 |
| Age | | Scalar | | 0.97 | | 0.08 | | 0.12 | | 0.00 | 0.00 |
| Sex | | Configural | | 0.97 | | 0.09 | | 0.11 | | - | - |
| Sex | | Metric | | 0.98 | | 0.08 | | 0.12 | | 0.01 | -0.01 |
| Sex | | Scalar | | 0.98 | | 0.08 | | 0.12 | | 0.00 | 0.00 |

Note: CFI values ≥ .95, RMSEA ≤ .08, and SRMR ≤ .08 generally indicate acceptable model fit.

**Figure S1.**

*Parent Academic Performance Ratings*

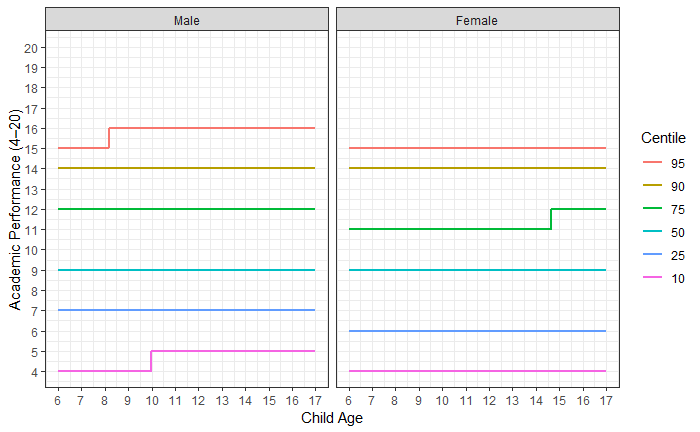

*Parent Social Performance Ratings*

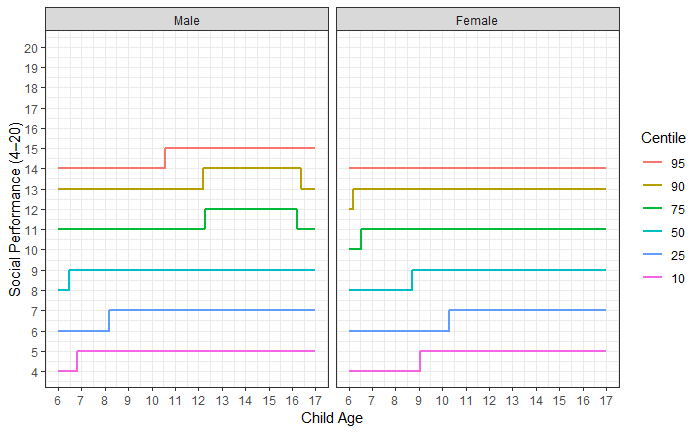

**Figure S2.**

*Adolescent Self-Report Academic Performance Ratings*

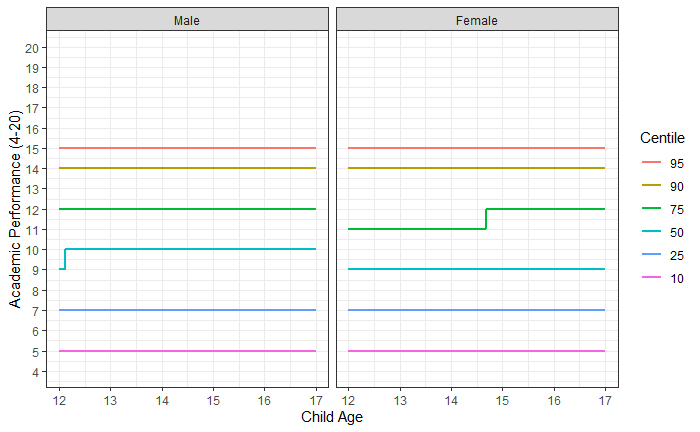

*Adolescent Self-Report Social Performance Ratings*

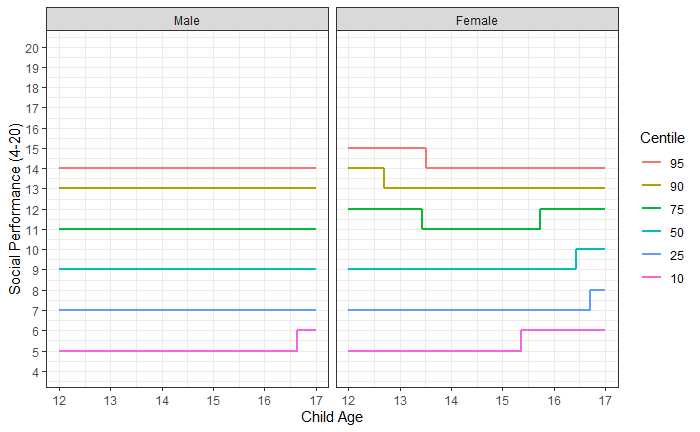

**Figure S3.** *Vanderbilt Parent Inattention Male*

Patient Name: _________________________________	Date: ________________

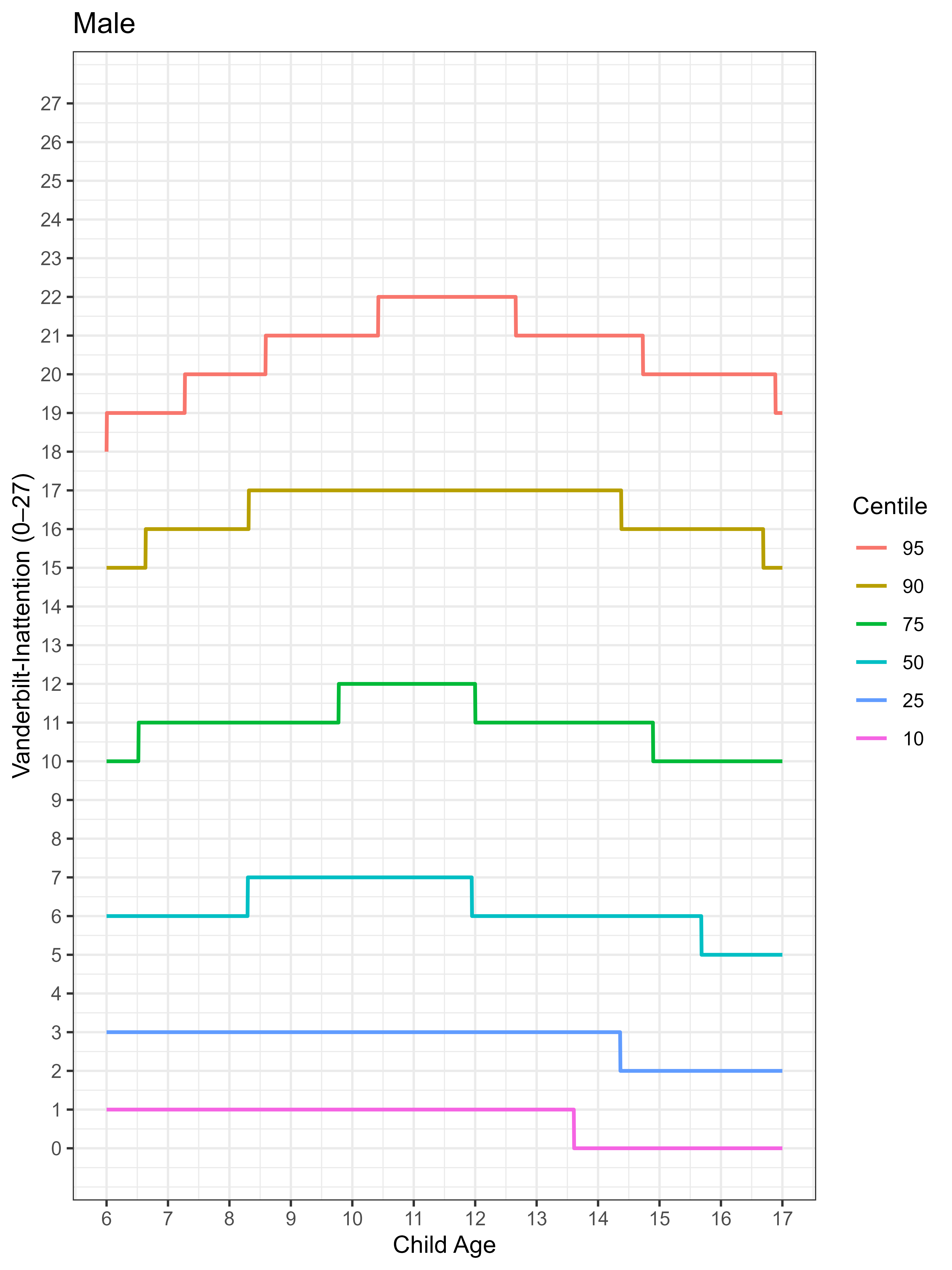

**Instructions to Clinician:** Calculate the Vanderbilt Inattention raw score by summing the 9 inattention items (range = 0-27). Locate the child’s age on the x-axis and raw score on the y-axis to determine the corresponding percentile. **Interpretation:** <75^th^ percentile = within the typical range; 75^th^ -89^th^ percentiles = elevated symptoms; ≥90^th^ percentile = substantial symptom elevation; ≥95^th^ percentile = very high levels of inattentive symptoms compared with same-age males. Percentiles are based on the normative reference sample and are intended to supplement, not replace, DSM-based symptom counts, impairment ratings, and clinical judgment.

**Figure S4.** *Vanderbilt Parent Inattention Female*

Patient Name: _________________________________	Date: ________________

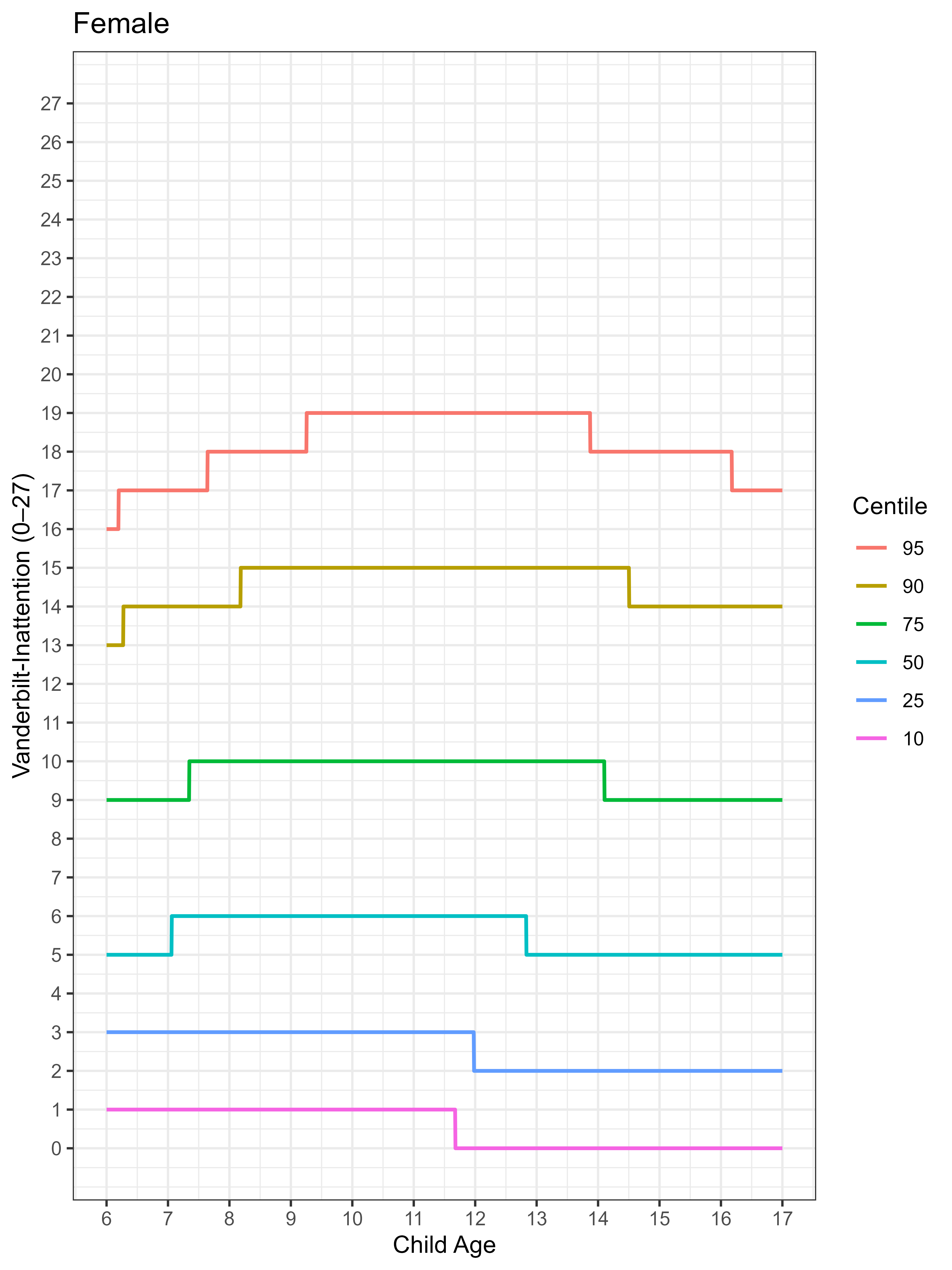

**Instructions to Clinician:** Calculate the Vanderbilt Inattention raw score by summing the 9 inattention items (range = 0-27). Locate the child’s age on the x-axis and raw score on the y-axis to determine the corresponding percentile. **Interpretation:** <75^th^ percentile = within the typical range; 75^th^ -89^th^ percentiles = elevated symptoms; ≥90^th^ percentile = substantial symptom elevation; ≥95^th^ percentile = very high levels of inattentive symptoms compared with same-age females. Percentiles are based on the normative reference sample and are intended to supplement, not replace, DSM-based symptom counts, impairment ratings, and clinical judgment.

**Figure S5.** *Vanderbilt Parent Hyperactivity/Impulsivity Male*

Patient Name: _________________________________	Date: ________________

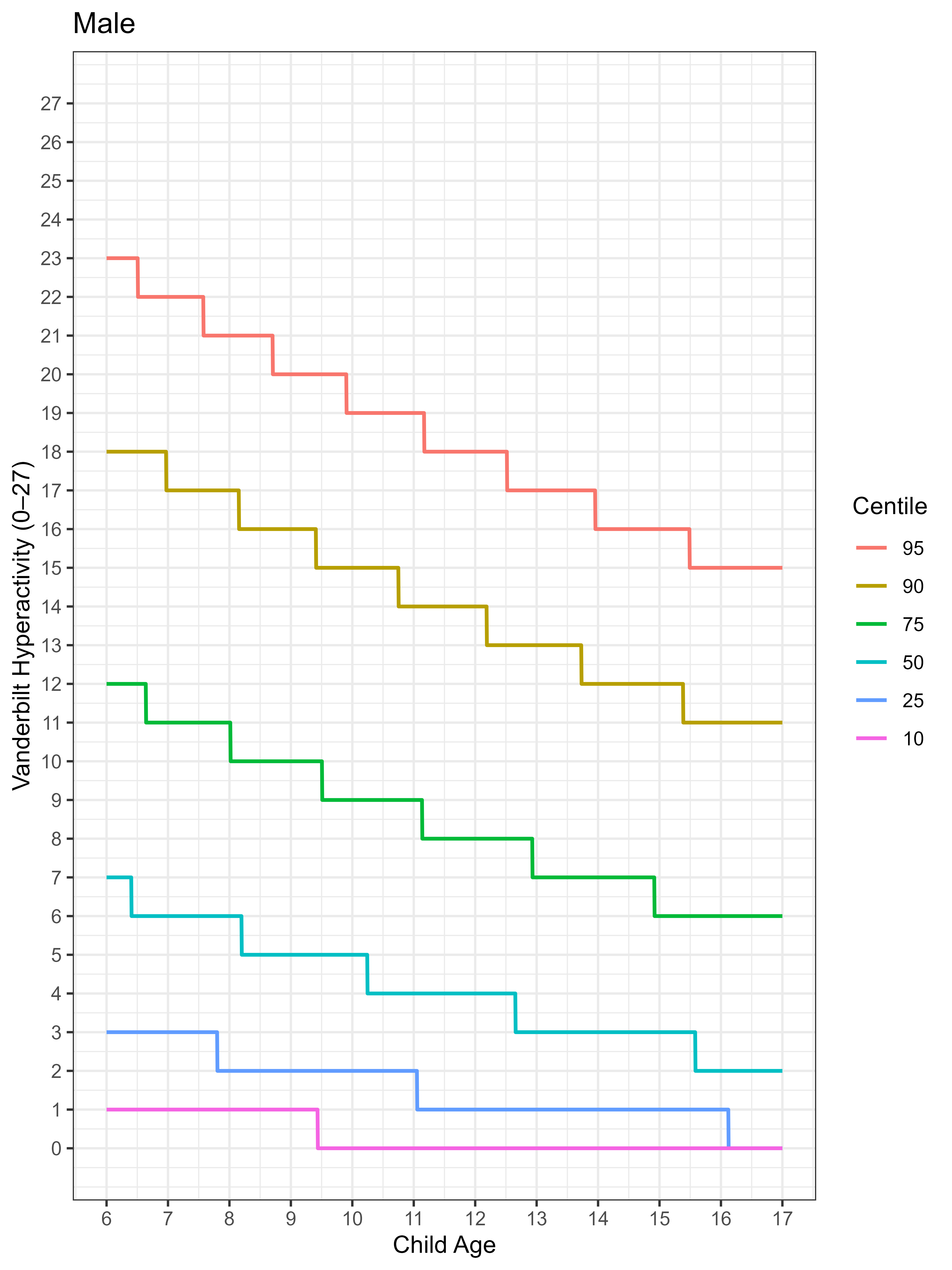

**Instructions to Clinician:** Calculate the Vanderbilt Hyperactivity/Impulsivity raw score by summing the 9 hyperactivity/impulsivity items (range = 0-27). Locate the child’s age on the x-axis and raw scores on the y-axis to determine the corresponding percentile. **Interpretation:** <75^th^ percentile = within typical range; 75^th^ -89^th^ percentiles = elevated symptoms; ≥90^th^ percentile = substantial symptom elevation; ≥95^th^ percentile = very high levels of hyperactivity/impulsivity symptoms compared with same-age males. Percentiles are based on the normative reference sample and are intended to supplement, not replace, DSM-based symptom counts, impairment ratings, and clinical judgment.

**Figure S6.** *Vanderbilt Parent Hyperactivity/Impulsivity Female*

Patient Name: _________________________________	Date: ________________

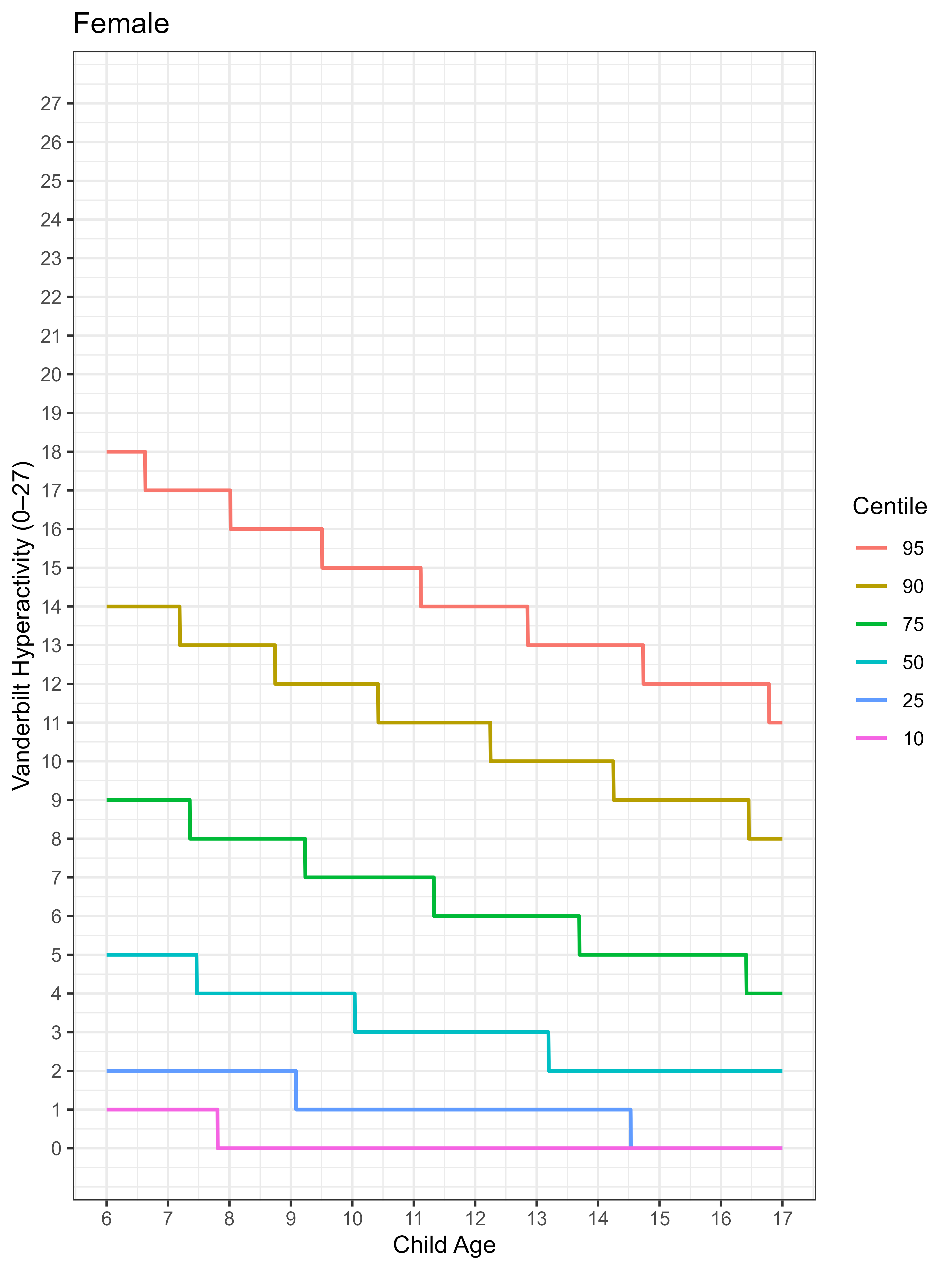

**Instructions to Clinician:** Calculate the Vanderbilt Hyperactivity/Impulsivity raw score by summing the 9 hyperactivity/impulsivity items (range = 0-27). Locate the child’s age on the x-axis and raw scores on the y-axis to determine the corresponding percentile. **Interpretation:** <75^th^ percentile = within typical range; 75^th^ -89^th^ percentiles = elevated symptoms; ≥90^th^ percentile = substantial symptom elevation; ≥95^th^ percentile = very high levels of hyperactivity/impulsivity symptoms compared with same-age females. Percentiles are based on the normative reference sample and are intended to supplement, not replace, DSM-based symptom counts, impairment ratings, and clinical judgment.
